# Multi-ancestry genome-wide association study reveals novel genetic signals for lung function decline

**DOI:** 10.1101/2024.11.25.24317794

**Authors:** Bonnie K. Patchen, Jingwen Zhang, Nathan Gaddis, Traci M. Bartz, Jing Chen, Catherine Debban, Hampton Leonard, Ngoc Quynh Nguyen, Jungkun Seo, Courtney Tern, Richard Allen, Dawn L. DeMeo, Myriam Fornage, Carl Melbourne, Melyssa Minto, Matthew Moll, George O’Connor, Tess Pottinger, Bruce M. Psaty, Stephen S. Rich, Jerome I. Rotter, Edwin K. Silverman, Jeran Stratford, R. Graham Barr, Michael H. Cho, Sina A. Gharib, Ani Manichaikul, Kari North, Elizabeth C. Oelsner, Eleanor M. Simonsick, Martin D. Tobin, Bing Yu, Seung Hoan Choi, Josee Dupuis, Patricia A. Cassano, Dana B. Hancock

## Abstract

**Rationale:** Accelerated decline in lung function contributes to the development of chronic respiratory disease. Despite evidence for a genetic component, few genetic associations with lung function decline have been identified.

**Objectives:** To evaluate genome-wide associations and putative downstream functionality of genetic variants with lung function decline in diverse general population cohorts.

**Methods:** We conducted genome-wide association study (GWAS) analyses of decline in the forced expiratory volume in the first second (FEV_1_), forced vital capacity (FVC), and their ratio (FEV_1_/FVC) in participants across six cohorts in the Cohorts for Heart and Aging Research in Genomic Epidemiology (CHARGE) consortium and the UK Biobank. Genotypes were imputed to TOPMed (CHARGE cohorts) or Haplotype Reference Consortium (HRC) (UK Biobank) reference panels, and GWAS analyses used generalized estimating equation models with robust standard error. Models were stratified by cohort, ancestry, and sex, and adjusted for important lung function confounders and genotype principal components. Results were combined in cross-ancestry and ancestry-specific meta-analyses. Selected top variants were tested for replication in two independent COPD-enriched cohorts.

**Measurements and Main Results:** Our discovery analyses included 52,056 self-reported White (N=44,988), Black (N=5,788), Hispanic (N=550), and Chinese American (N=730) participants with a mean of 2.3 spirometry measurements and 8.6 years of follow-up. Functional mapping of GWAS meta-analysis results identified 361 distinct genome-wide significant (p<5E-08) variants in one or more of the FEV_1_, FVC, and FEV_1_/FVC decline phenotypes, which overlapped with previously reported genetic signals for several related pulmonary traits. Of these, 8 variants, or 20.5% of the variant set available for replication testing, were nominally associated (p<0.05) with at least one decline phenotype in COPD-enriched cohorts (White [N=4,778] and Black [N=1,118]). Using the GWAS results, gene-level analysis implicated 38 genes, including eight (*XIRP2*, *GRIN2D*, *SATB1*, *MARCHF4*, *SIPA1L2*, *ANO5*, *H2BC10*, and *FAF2*) with consistent associations across ancestries or decline phenotypes. Annotation class analysis revealed significant enrichment of several regulatory processes for corticosteroid biosynthesis and metabolism. Drug repurposing analysis identified 43 approved compounds targeting eight of the implicated 38 genes.

**Conclusions:** Our multi-ancestry GWAS meta-analyses identified numerous genetic loci associated with lung function decline. These findings contribute knowledge to the genetic architecture of lung function decline, provide evidence for a role of endogenous corticosteroids in the etiology of lung function decline, and identify drug targets that merit further study for potential repurposing to slow lung function decline and treat lung disease.

## INTRODUCTION

Lung function increases through childhood, plateaus in early adulthood, then declines with age. A faster rate or accelerated decline in lung function is associated with chronic lung disease, including chronic obstructive pulmonary disease (COPD), a leading cause of death worldwide. Inflammation from smoking and other environmental exposures contributes to accelerated lung function decline and development of chronic lung disease, but not all smokers develop lung disease and non-smokers can develop chronic lung disease as well.

Genetic susceptibility also plays a role in lung function. Genome-wide association studies (GWAS) have identified hundreds of genetic loci associated with cross-sectional lung function, with heritability estimates ranging from 20%–40% for forced expiratory volume in the first second (FEV_1_), forced vital capacity (FVC), and their ratio (FEV_1_/FVC).^3–5^ Heritability analyses suggest that there is also a genetic component to change in lung function over time, with estimates ranging from 5%–18% across lung function phenotypes.^6–8^ However, GWAS evaluating lung function decline are limited, and few genetic loci associated with lung function decline at genome-wide significance have been identified.^9–11^ A prior GWAS meta-analysis of decline in FEV_1_ that included over 27,000 European ancestry (EA) participants from 14 cohorts identified one locus, *ME3* on chromosome 11, that was associated with FEV_1_ decline at genome-wide significance.^12^ Later, a GWAS of lung function decline in around 6,500 Korean participants identified a variant near *CEP164*, also on chromosome 11, associated with annual change in FEV_1_/FVC.^7^ Another GWAS in an independent cohort of 6,800 Korean participants identified one variant in *SLC6A1* (chromosome 3) associated with decline in both FEV_1_ and FVC.^8^ Other GWAS of lung function decline with smaller sample sizes or shorter follow-up periods have not yielded genome-wide significant associations.^10,11^ Larger GWAS with increased power are needed to identify robust genetic signals for lung function decline.

Here, we conducted the largest GWAS of lung function decline to date (total N = 52,056), included multiple ancestries, and achieved more coverage of the genome via newer imputation platforms to improve resolution of genetic signals for lung function decline. We identified novel variants associated with decline in FEV_1_, FVC, and FEV_1_/FVC at genome-wide significance and connected these variants to their corresponding genes, biologically relevant pathways, and identified drug targets that could be repurposed to slow decline and treat lung disease. These findings shed light on the genetic contribution to lung function decline.

## Methods

### Study populations and measurements

Our GWAS meta-analyses included six US-based studies from the Cohorts for Heart and Aging Research in Genomic Epidemiology (CHARGE) Consortium—Atherosclerosis Risk in Communities (ARIC), Coronary Artery Risk Development in Young Adults (CARDIA), Cardiovascular Health Study (CHS), Framingham Heart Study (FHS), Health, Aging and Body Composition Study (HABC), and Multi-Ethnic Study of Atherosclerosis (MESA)—and the UK Biobank. All studies included White/EA participants, five studies included Black/African ancestry (AA) participants, and one study included Hispanic and Chinese American participants. Local ethics committees and Institutional Review Boards reviewed protocols in each study. All study participants provided written informed consent.

FEV_1_ and FVC were measured by spirometry at baseline and follow-up visits following American Thoracic Society or European Respiratory Society recommendations current at time of assessment. FEV_1_ and FVC measurements passing quality control criteria for acceptability were included. The ratio of FEV_1_ to FVC (FEV_1_/FVC) was calculated when both FEV_1_ and FVC passed acceptability criteria. Pre-bronchodilator measures were used for all discovery studies.

Genotyping in each study was performed as previously described and following standard quality control measures.^11^ Imputation was based on the Trans-Omics for Precision Medicine (TOPMed; for U.S.-based CHARGE studies) or Haplotype Reference Consortium (HRC; for UK Biobank) reference panels. Within each study, variants with imputation values <0.3 were excluded. See the online supplement for additional details on contributing studies and their spirometry measurements and genotyping and imputation methods.

### Statistical analysis

Figure 1 presents an overview of the analysis scheme. We performed genome-wide analyses for decline in FEV_1_, FVC, and FEV_1_/FVC separately in each study following a standardized analysis plan. The NHLBI Pooled Cohorts harmonized spirometry data in each cohort were used where possible (CHARGE studies).^13^ We estimated genome-wide variant associations with lung function decline using generalized estimating equations with robust standard error (GEE) and unstructured correlation structure. In all models, repeated measurements of FEV_1_, FVC, or FEV_1_/FVC were regressed on variant, elapsed time since first lung function measurement, and the variant × elapsed time multiplicative interaction term. We focused on the variant x time interaction term to identify variants associated with lung function decline. The GEE model was selected based on its robustness for testing interaction terms in genetic association studies even when the interaction variable is mis-specified.^14^ Analyses were performed in R studio using the geepack R package.^15^

**Figure 1.**
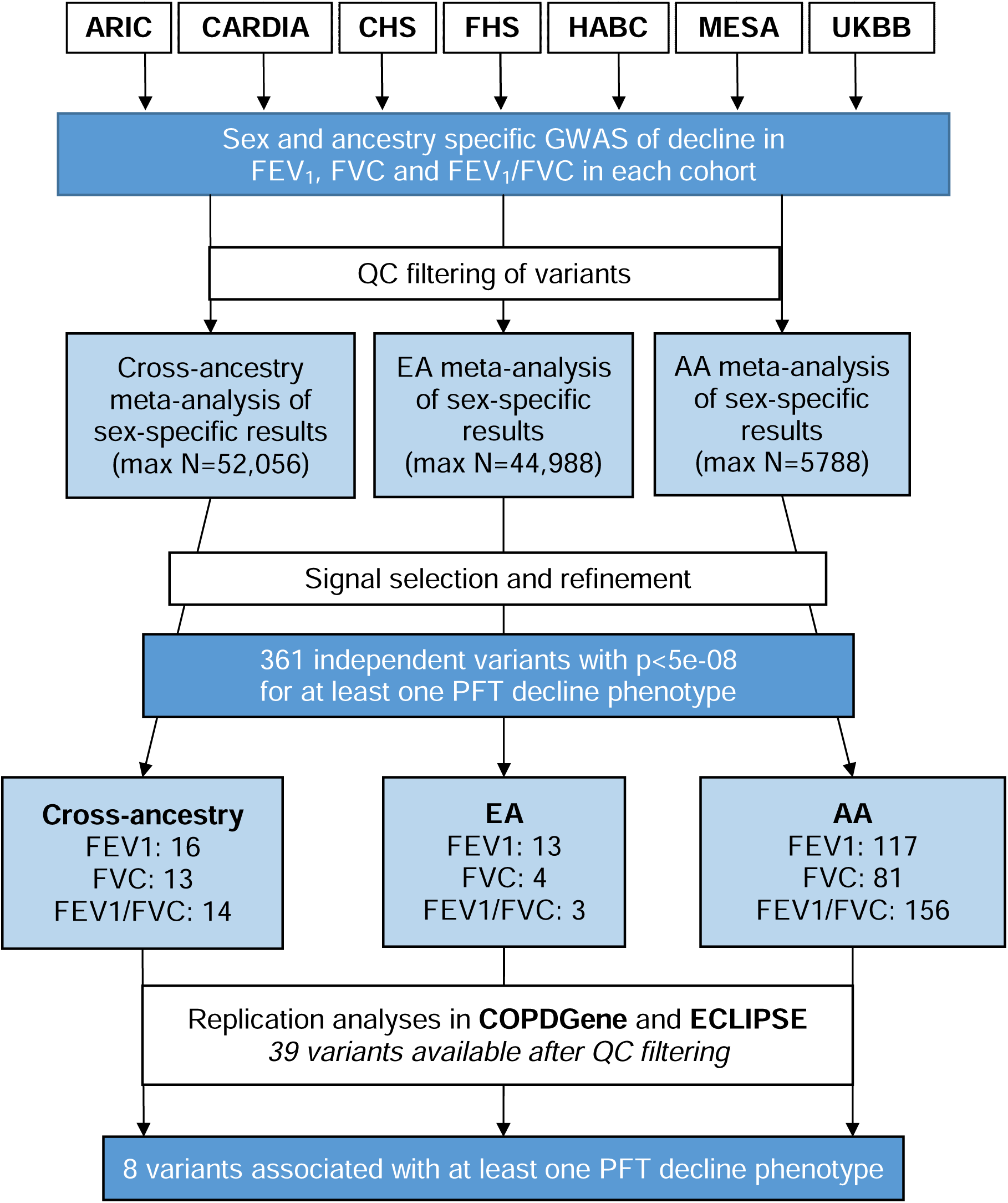
Study Design. Sex and ancestry-stratified GWAS analyses were performed separately in each cohort following a uniform analysis plan and summarized with inverse-variance weighted meta-analysis. Distinct variants with p-values passing genome-wide significance thresholds (p < 5E-08) were considered significant. Significant variants were tested for replication in two COPD-enriched cohorts following the discovery analysis plan. Replication was declared for variants with p-values passing the Bonferroni-adjusted significance threshold (0.05/361 distinct variants).

We stratified models by self-reported ancestry and sex and adjusted them for baseline age, mean centered baseline age^2^, time-varying height, mean centered height^2^, weight (FVC only), baseline smoking status (current, former, never), current smoking at each spirometry measurement (yes/no), baseline smoking pack-years, cigarettes per day at each spirometry visit, study site (for multi-site cohorts), and genotype principal components. Mean centering of squared terms was done to minimize collinearity. Weight was included in the FVC model only based on its stronger association with a restrictive (FVC) versus obstructive (FEV_1_) phenotype. For cohorts that had a change in spirometry equipment during follow-up we also included a variable indicating whether lung function measurements occurred before or after the spirometer change. See the online supplement for additional details of the statistical models. To remove extreme values driven by small sample sizes, prior to meta-analysis, we filtered sex- and ancestry-specific results from each study to exclude variants present in <30 participants, variants with minor allele counts <20, and variants with effect sizes <−50 or >50 mL/year.

Meta-analyses of ancestry- and sex-specific results from each study were performed with METAL using the inverse-variance based fixed effects method. The primary cross-ancestry analyses combined all ancestry- and sex-specific results for each lung function decline phenotype. Secondary analyses included ancestry-specific meta-analyses for EA and AA results; sample sizes for the other ancestries were too small to conduct ancestry-specific meta-analyses. We further filtered meta-analysis results on total minor allele count <200, variants present in <25% of the maximum sample size, or variants present in fewer than two of the sex- and ancestry-specific cohort results. For the cross-ancestry analyses, due to the limited sample sizes for Hispanic and Chinese American analyses we required that variants also be present in EA or AA analyses.

### Signal selection

To identify distinct significant variants, we processed the meta-analysis results using the Functional Mapping and Annotation (FUMA) SNP2GENE function.^12^ Lead and candidate variants were selected using the FUMA SNP2GENE default parameters (significance threshold of p<5E-08; r^2^ thresholds to define lead and candidate variants of 0.1 and 0.6; 1000G Phase3 ALL reference panel; maximum distance of 250 kb between LD blocks to merge into a locus). More stringent significance thresholds may be needed for genomes with less linkage disequilibrium, therefore we also report the number of variants significant at 2.5E-08 for the cross-ancestry and AA analyses.^16,17^ Decline-associated variants and variants within ±500 kb were evaluated for associations with relevant spirometry or lung disease phenotypes in the GWAS catalogue and Phenoscanner databases.^13–15^ We used the ±500 kb window for queries rather than an LD threshold because the majority of variants had limited information on LD as they were were not in the 1000 Genomes reference panel. All queries were performed in R studio 3.4 using the LDlinkR and Phenoscanner R packages.^16,17^ We performed additional lookups for variants in the GWAS catalogue reported to be associated with spirometry (FEV_1_, FVC) and lung disease (asthma, COPD, emphysema, chronic bronchitis).

### Replication of lung function decline-associated variants in COPD-enriched populations

We further evaluated variants associated with lung function decline in the cross-ancestry or ancestry-specific analyses in White (N=4,778) and Black (N=1,118) participants in two cohort studies enriched for participants with COPD: the Genetic Epidemiology of COPD (COPDGene)^18^ and the Evaluation of COPD Longitudinally to Identify Predictive Surrogate End-points (ECLIPSE).^19^ Following the same analysis plan as the primary analyses described above, we estimated associations of variants with decline in FEV_1_, FVC and FEV_1_/FVC, and performed meta-analyses of sex- and ancestry- specific results for each decline phenotype. Because prevalent COPD and COPD severity may influence longitudinal lung function, we repeated the analyses adjusting for COPD status and disease stage (no COPD/preserved ratio impaired spirometry [PRISm]/GOLD status 1/2/3/4).^18–20^ Only variants passing QC filtering criteria (MAC>20 in at least two sex and ancestry strata) with genomic position and reference/alternate alleles matching those identified in the discovery analyses were tested. We used Bonferroni correction for the number of variants tested for assessing stringent statistical significance, along with a nominal significance threshold of p<0.05 for declaring suggestive replication.

### Heritability and genetic correlation with relevant pulmonary traits

Heritability of lung function decline and genetic correlation of decline phenotypes with relevant pulmonary traits were estimated in the FHS cohort (N=3,571 EA participants) using SOLAR. We limited this analysis to FHS for three reasons: (1) FHS is family-based so we could use family relationships to estimate heritability, (2) FHS is an optimal cohort for heritability calculations because of the number and time separation in the set of repeated measurements of lung function, and (3) we had access to individual-level data in FHS which was critical because our GWAS evaluated lung function decline with an interaction term and to our knowledge there are currently no validated methods for estimating heritability of interaction terms that use summary statistics. We evaluated decline phenotypes with evidence of significant heritability (p<0.05) for genetic correlation with baseline FEV_1_ and FVC, COPD at baseline, COPD at last observation, carbon monoxide diffusion capacity (DLCO), asthma, immunoglobulin E (IgE), emphysema, and interstitial lung abnormality (ILA). All heritability and genetic correlation analyses accounted for sex, baseline age, and ever/never smoking status.

### Gene-based tests and enrichment analyses

Gene-based tests were performed in MAGMA for each set of meta-analysis results using the SNP-wise mean, SNP-wise top, and SNP-wise multi (aggregate of SNP-wise mean and SNP-wise top) methods.^21^ Genes with z-scores significant at Bonferroni-corrected p-value thresholds (p<0.05/18,156 genes) for one or more of the MAGMA methods were considered significant.

Gene expression in lung tissue was evaluated using the GTEx platform.^22^ We evaluated pulmonary traits associated with variants mapped to decline-associated genes reported in the GWAS catalogue and Phenoscanner databases using the LDlinkR and Phenoscanner R packages.^23–27^

Enrichment analyses were based on positional mapping of candidate variants to genes and genomic loci using FUMA. We evaluated the enrichment of pre-defined gene sets obtained from the Molecular Signatures Database, WikiPathways and reported genes from the GWAS catalogue with the FUMA GENE2FUNC function.^28^ Multiple testing correction was performed separately for each gene-set category. Gene sets with Benjamini-Hochberg false discovery rate adjusted p-values <0.05 were considered enriched. Pathway and disease enrichment patterns were explored for FUMA-defined genomic loci by annotation class analysis using the Genomic Regions Enrichment of Annotations Tool (GREAT).^29,30^ Pathway and disease terms with false discovery rate–adjusted p-values <0.05 for binomial or hypergeometric tests were declared significant.

### Drug repurposing analyses

Protein coding genes that were significant for at least one decline phenotype from the MAGMA analysis were labeled as priority genes that we evaluated to identify candidates for drug repurposing studies. We identified existing pharmacotherapies targeting prioritized genes by querying four large drug-related resources, including Pharos, a web interface for the Target Central Resource Database (http://juniper.health.unm.edu/tcrd/),^31,32^ Open Targets,^33^ Therapeutic Target Database,^34^ and DrugBank.^35^ A cross-resource summary was generated for expert review that includes level of clinical development (preclinical, in clinical trials, approved), gene target selectivity (total number of targets), and other relevant metrics (approved uses, safety).

## Results

### Participants

Our analyses included 52,056 self-reported White (N=44,988), Black (N=5788), Hispanic (N=550), and Chinese American (N=730) participants (Figure 1, Table 1). Fifty eight percent were never smokers, 32% were former smokers, and 10% were current smokers. Participants were followed for a mean of 8.6 years with a mean of 2.3 spirometry measurements over the follow-up period. Cohort- and race and ethnicity-specific means for years of follow-up and number of spirometry measurements ranged from 4.2 18.7 and 1.7 4.6, respectively. Participant characteristics by self-reported race and ethnicity are shown in Table 1, and characteristics for each study are presented in the online supplement (Table S1).

**Table 1.** Participant Characteristics.

| N | Total | European ancestry | African ancestry | Hispanic | Chinese ancestry |
| --- | --- | --- | --- | --- | --- |
|  | 52,056 | 44,988 | 5,788 | 730 | 550 |
| Cohort, N (%) |  |  |  |  |  |
| ARIC | 11,091 (21.3) | 8,704 (19.3) | 2,387 (41.2) | — | — |
| CARDIA | 2,550 (4.9) | 1,611 (3.6) | 939 (16.2) | — | — |
| CHS | 3,663 (7.0) | 3,085 (6.9) | 578 (10.0) | — | — |
| FHS | 2,013 (3.9) | 2,013 (4.5) | — | — | — |
| HABC | 2,464 (4.7) | 1,505 (3.3) | 959 (16.6) | — | — |
| MESA | 3,876 (7.4) | 1,671 (3.7) | 925 (16.0) | 730 (100.0) | 550 (100.0) |
| UKBB | 26,399 (50.7) | 26,399 (58.7) | — | — | — |
| Female, N (%) | 28,455 (54.7) | 24,718 (54.9) | 3,348 (57.8) | 389 (53.5) | 270 (49.1) |
| Age, yr | 56.5 (11.9) | 56.2 (10.9) | 56.5 (17.5) | 66.4 (10.3) | 66.0 (9.7) |
| Height, cm | 168.3 (9.3) | 168.6 (9.2) | 167.3 (9.3) | 161.3 (9.2) | 161.5 (8.7) |
| Weight, kg | 76.4 (15.7) | 76.0 (15.4) | 80.6 (17.4) | 77.2 (15.7) | 63.0 (11.0) |
| Current smoking, N (%) | 5,390 (10.4) | 4,180 (9.3) | 1,128 (19.5) | 58 (7.9) | 24 (4.4) |
| Former smoking, N (%) | 1,6629 (31.9) | 1,4315 (31.8) | 1,843 (31.8) | 330 (45.2) | 141 (25.6) |
| Baseline FEV<sub>1</sub>, mL | 2,789 (814) | 2,860 (799) | 2,351 (773) | 2,328 (720) | 2,189 (653) |
| Baseline FVC, mL | 3,696 (1016) | 3,798 (994) | 3,060 (916) | 3,059 (897) | 2,915 (841) |
| Baseline FEV<sub>1</sub>/FVC, % | 75.4 (7.5) | 75.2 (7.2) | 76.7 (9.0) | 76.1 (8.0) | 75.2 (7.4) |
| N PFT measurements | 2.3 (1.0) | 2.3 (0.9) | 2.3 (1.9) | 1.7 (0.8) | 1.8 (0.8) |

| **N** | **Total** | **European ancestry** | **African ancestry** | **Hispanic** | **Chinese ancestry** |
| --- | --- | --- | --- | --- | --- |
|  | **52,056** | **44,988** | **5,788** | **730** | **550** |
| Total follow-up, years | 8.6 (6.3) | 8.7 (6.0) | 8.3 (8.2) | 4.2 (4.9) | 5.5 (5.2) |
| Time between pulmonary function test visits, years | 7.3 (5.2) | 7.1 (5.0) | 8.0 (7.1) | 7.6 (3.2) | 8.2 (3.2) |

Values are presented as mean (SD) unless otherwise specified.

### GWAS meta-analyses identify robust associations with lung function decline

Genome-wide meta-analyses captured up to 11.8 million genotyped and TOPMed- or HRC-imputed variants. Genomic inflation values (λ_gc_) from cohort-specific results ranged from 0.57 to 1.30 after QC filtering, and results with λ_gc_ > 1 were adjusted prior to meta-analysis using the genomic control function in METAL. After combining results, we found little evidence for genomic inflation in the cross-ancestry (λ_gc_= 1.03 for FEV_1_ and 1.02 for both FVC and FEV_1_/FVC), EA (λ_gc_ =1.03 for FEV_1_, 1.01 for FVC and 1.02 for FVC), or AA (λ_gc_ =1.08 for FEV_1_, 1.05 for FVC and 1.03 for FEV_1_/FVC) meta-analyses (Figure S1).

Cross-ancestry meta-analyses identified 39 variants representing distinct genomic regions (R^2^<0.1) associated with lung function decline at p<5E-08 (Figure 2, Table S2). Of these, 12 variants were associated with FEV_1_, nine were associated with FVC, four were associated with both FEV_1_ and FVC, and 14 were associated with FEV_1_/FVC. These 39 variants represent novel signals for longitudinal decline in lung function, and most of the variants (36 of 39) had minor allele frequencies <0.05 in the study population (Figure S2). 32 of the 39 variants were significant at the more stringent threshold of p<2.5E-08 recommended Ancestry-specific meta-analyses identified 20 distinct variants for EA (13 for FEV_1_, four for FVC, and three for FEV_1_/FVC; Figure S3) and 307 distinct variants for AA (79 for FEV_1_, 45 for FVC, and 138 for FEV_1_/FVC, 9 for both FEV_1_ and FEV_1_/FVC, 7 for both FVC and FEV_1_/FVC, and two for all three phenotypes; Figure S4). All 20 variants significant in the EA analyses had minor allele frequencies <0.05, whereas most variants significant in the AA analyses (224 of 307) had minor allele frequencies ≥0.05 (Figure S2). 277 of the variants identified in the AA analyses were significant at the more stringent threshold of p<2.5E-08.^17^ Of note, a majority (87%) of the variants identified in the AA analyses were present (i.e. passed our QC filtering criteria for MAC) only in the ARIC cohort, which we attribute to ARIC having largest sample size for AA participants (∼41% of the total AAs; total N=2,387, including 1,482 females and 905 males).

**Figure 2.**
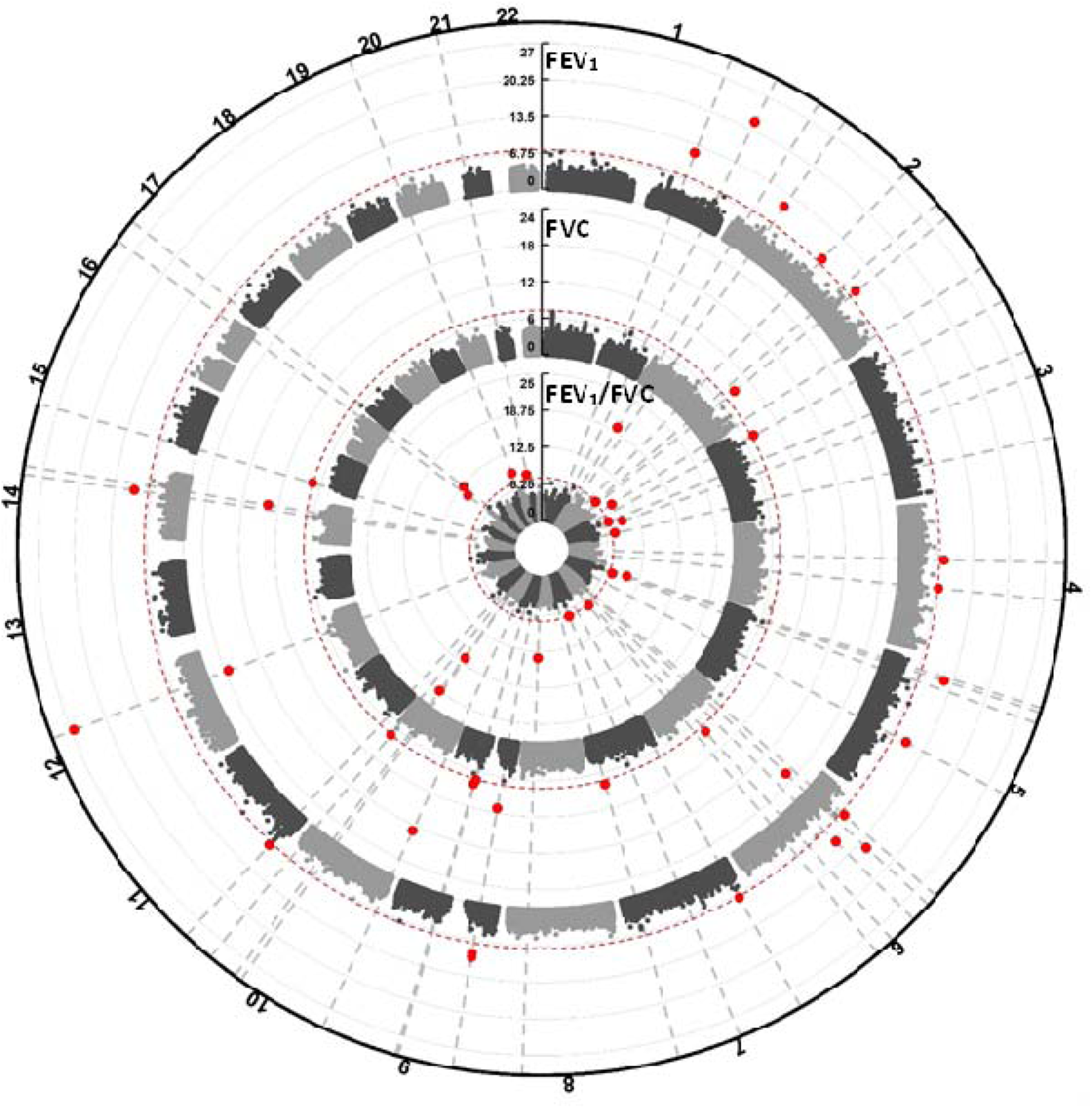
Circular Manhattan Plot of Cross-Ancestry Results. Genome-wide results for decline in FEV_1_ (outer circle), FVC (middle circle), and FEV_1_/FVC (inner circle) from the cross-ancestry analyses. Dotted red lines denotes the genome-wide significance threshold of p = 5E-08. Red circles represent variants passing genome-wide significance.

Eleven variants significant in the cross-ancestry meta-analyses were also significant or within 500 kb of variants significant in the EA- or AA-specific meta-analyses (Table 2). One variant significant only in the AA analyses, rs32137 in the *CTNND2* gene on chromosome 5, is within ±500 kb of a variant previously reported for nominal association (p=6E-06) with FEV_1_ decline in a GWAS of asthmatics.^8^ None of the other variants identified in either the EA or AA analyses have previously been associated with lung function decline.

**Table 2.** Cross-ancestry variants significant or within 500 kb of variants significant in the European or African ancestry analyses.

| **rsID** | **Chromosome**<br>**:position**<br>**(hg38)** | **Reference/**<br>**alternate alleles** | **Closest Gene** | **\*N**<br>**Studies** | **\*Total**<br>**Sample**<br>**size** | **\*MAF** | **\*Phenotype** | **\*Effect**<br>**Estimate** | **\*P-value** |
| --- | --- | --- | --- | --- | --- | --- | --- | --- | --- |
| rs114110050 | 1:181557627 | G/C | *CACNA1E* | 9 | 41303 | 0.02 | FEV<sub>1</sub> | −6.07 | 1.0E-12 |
| rs561006775 | 2:43650017 | AG/A | *PLEKHH2* | 7 | 15231 | 0.20 | FEV<sub>1</sub> | 7.48 | 6.3E-12 |
| rs139399765 | 3:20815668 | C/T | *RNU6-815P* | 7 | 39318 | 0.04 | FVC | −4.82 | 2.2E-08 |
| rs149963074 | 3:99726132 | C/T | *COL8A1* | 9 | 16903 | 0.02 | RATIO | 0.13 | 2.1E-10 |
| rs73875952 | 3:161893323 | G/C | *RP11-774I5.1* | 6 | 14219 | 0.01 | RATIO | −0.10 | 1.4E-08 |
| rs111351387 | 4:71826274 | G/T | *RP11-545L5.1* | 14 | 20927 | 0.06 | FEV<sub>1</sub> | 3.83 | 6.3E-09 |
| rs559126463 | 6:55623623 | G/GATATATAC | *HMGCLL1* | 11 | 18621 | 0.04 | FEV<sub>1</sub> | 4.64 | 5.2E-11 |
| rs78349759 | 6:83786677 | C/T | *RP3-393K13.1* | 9 | 41348 | 0.01 | FVC | 5.64 | 4.2E-09 |
| rs150709492 | 7:8801178 | C/T | *NXPH1* | 15 | 46421 | 0.02 | FEV<sub>1</sub> | −3.81 | 2.9E-08 |
| rs2921778 | 8:116877202 | G/T | *RAD21-AS1* | 13 | 16627 | 0.03 | RATIO | 0.10 | 2.2E-14 |
| rs540140520 | 9:78892132 | AC/A | *KRT18P24* | 6 | 27725 | 0.01 | FVC | −6.05 | 6.7E-09 |

\*N studies, total sample size, alternate allele frequency, decline phenotype, effect estimate, and p-value are from cross-ancestry analysis.

### Genetic signals for lung function decline suggest replicability in independent COPD-enriched populations

Of the 361 distinct variants identified in our primary analysis, a limited set of 39 were available (passing QC criteria and aligned for genomic position and reference/alternative alleles) for replication testing in COPDGene and/or ECLIPSE. This included 28 variants for the EA meta-analysis, 12 variants for the AA meta-analysis and 4 variants for the cross-ancestry meta-analysis. No variants surpassed Bonferroni correction for the 39 variants tested for replication (p<0.0013 [p<0.05/39)]). Six variants (15.4% of those tested) were nominally significantly associated (p<0.05) with at least one decline phenotype in the primary analysis (Table 3).

**Table 3.** Decline variants with nominal evidence (p<0.05) for decline associations in COPD-enriched Cohorts.

| rsID | Chromosome:<br>position | Association in Discovery Cohorts |  |  |  |  |  | Association in Replication Cohorts |  |  |  |  |  |
| --- | --- | --- | --- | --- | --- | --- | --- | --- | --- | --- | --- | --- | --- |
|  |  | Ance<br>stry | Pheno | EA | EAF | Effect<br>mL or<br>% per<br>year | P-value | Ance<br>stry | Pheno | EA | EAF | Effect<br>mL or<br>% per<br>year | P-value |
| Associations seen in primary replication analysis |  |  |  |  |  |  |  |  |  |  |  |  |  |
| rs57862396 | 9:33245969 | AA | RATIO | T | 0.05 | 0.48 | 3.94E-08 | EA | RATIO | T | 0.03 | 0.13 | 0.01124 |
| rs876508 | 13:45863985 | AA | RATIO | T | 0.03 | 0.14 | 3.89E-08 | EA | RATIO | T | 0.06 | -0.14 | 5.28E-04 |
| rs6814777 | 4:142766606 | AA | RATIO | A | 0.85 | 0.37 | 2.74E-14 | EA | FVC | A | 0.97 | 37.5 | 0.03264 |
| rs113816940 | 7:55634105 | EA | FEV | A | 0.02 | 3.82 | 3.88E-09 | EA | FVC | A | 0.02 | -7.75 | 0.04785 |
| rs200755764 | 11:11868692 | ALL | FEV | G | 0.03 | 4.92 | 1.12E-08 | AA | FVC | G | 0.05 | -10.7 | 0.02936 |
| rs74154586* | 10:103676221 | AA | FVC | T* | 0.04 | 20.6 | 2.60E-25 | AA | RATIO | A* | 0.05 | -0.23 | 0.02272 |
| Additional associations seen in replication analysis adjusting for COPD status and disease stage |  |  |  |  |  |  |  |  |  |  |  |  |  |
| rs12445073 | 16:86705263 | AA | RATIO | T | 0.09 | 0.19 | 1.32E-08 | EA | FVC | T | 0.04 | 41.9 | 0.02729 |
| rs13391412 | 2:116850979 | AA | FVC | T | 0.04 | -20.8 | 2.76E-08 | EA | RATIO | T | 0.02 | -0.21 | 0.02897 |
| rs200755764 | 11:11868692 | ALL | FEV | G | 0.03 | 4.92 | 1.12E-08 | AA | FEV | G | 0.05 | -7.3045 | 0.04762 |

Pheno: pulmonary function decline phenotype; EA: effect allele; EAF: effect allele frequency
\*rs74154586 is a triallelic variant with C as the reference allele and A and T both being alternate alleles. The C\_T variant was identified in the discovery cohorts but only the C\_A variant was present in the replication cohorts.

Results were similar in models adjusted for COPD status and disease stage (correlation coefficient [corr] = 0.90, 95% CI = 0.86 to 0.93), with three additional associations (two additional variants) reaching nominal significance thresholds in this analysis (Table 3). Only one variant, rs57862396 on chromosome 9, replicated for the same phenotype (FEV1/FVC) and with the same direction of effect. For this variant, the T (alternate) allele was associated with a slower rate or attenuation in FEV1/FVC decline of 0.48% per year in the discovery cohorts and with an attenuation of 0.13% per year in the replication cohorts (p-discovery=3.939E-08, p- replication=0.011). Two other variants showed associations with the same phenotype across the discovery and replication analyses (rs876508 on chromosome 13 for FEV1/FVC and rs200755764 on chromosome 11 for FEV1), but with opposite directions of effect. The other nominally significant associations were observed for different phenotypes between the discovery and replication analyses, including three with the same direction of effect and three with the opposite direction of effect. P-values for each ancestry and phenotype for all variants available (passing QC criteria) for the replication analysis are presented in Table S4.

### Genetic signals for lung function decline overlap with signals for relevant pulmonary phenotypes

Evaluation of decline-associated variants for associations with relevant pulmonary traits in the GWAS catalogue and Phenoscanner databases provided evidence for pleiotropy across pulmonary phenotypes (Figure 3, Tables S5 and S6). Of the 361 distinct decline-associated variants (defined by R^2^<0.1) identified in the cross-ancestry or ancestry-specific analyses, 302 variants were ±500 kb of variants associated with at least one other pulmonary phenotype, including cross-sectional lung function (58 variants); COPD (27 variants); asthma/allergic disease (85 variants); emphysema or chronic bronchitis (155 variants); pulmonary fibrosis, interstitial lung disease, or other unspecified respiratory disease (169 variants); and death from respiratory disease (48 variants) (Figure 3). There was also a large degree of overlap across these variant sets: 103 decline variants were ±500 kb of variants associated with two other pulmonary traits; 39 were ±500 kb of variants associated with three other pulmonary traits; 17 were ±500 kb of variants associated with four other pulmonary traits; and two variants, rs569142166 in the *RAB31* gene on chromosome 18 and rs561379073 near the *LINCO1139* gene on chromosome 1, were ±500 kb of variants associated with five of the six pulmonary traits evaluated (Figure 3).

**Figure 3.**
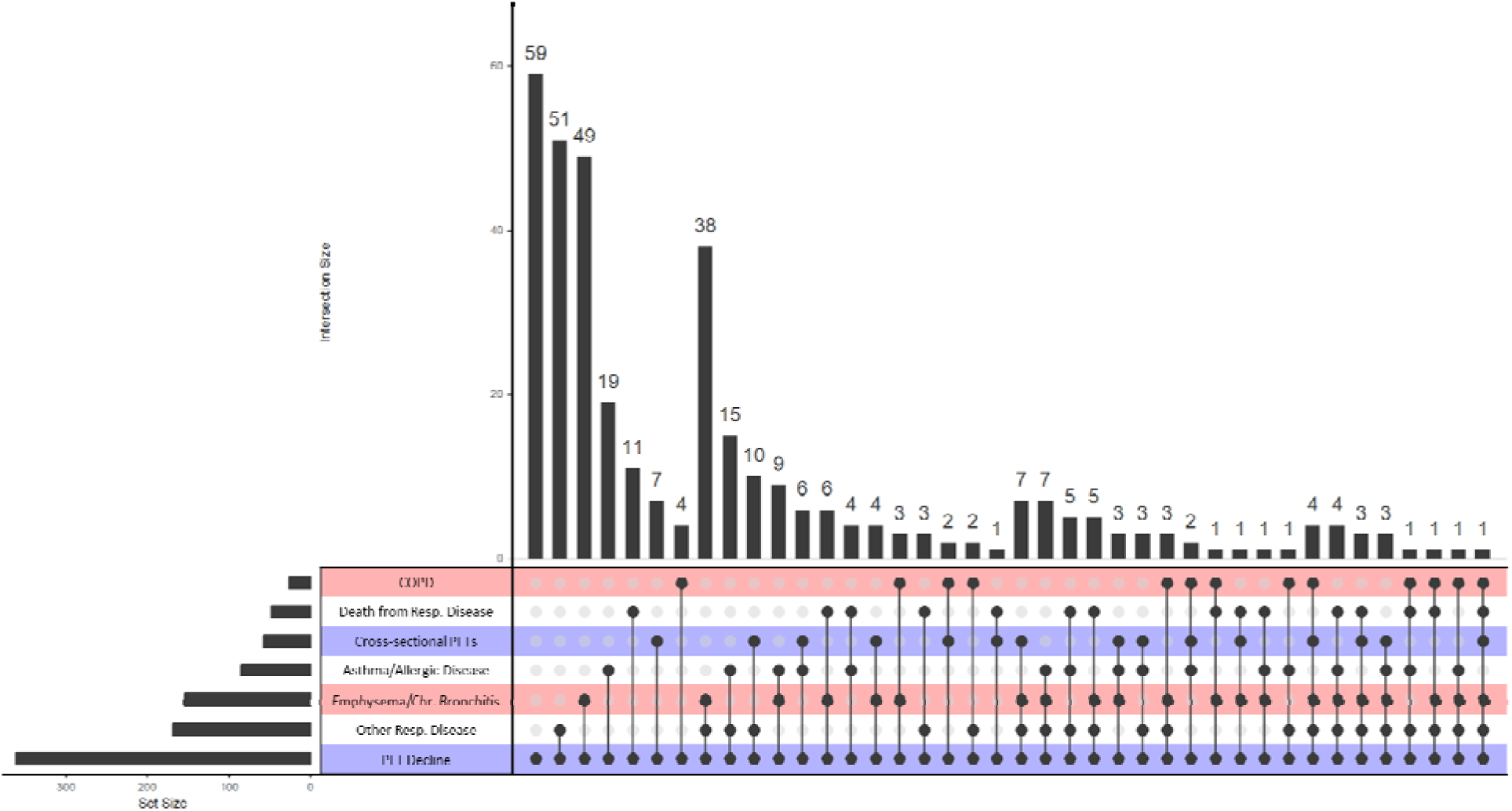
Upset Plot Showing Overlap of Lung Function Decline-Associated Variants with Previously Reported Signals for Relevant Pulmonary Phenotypes. Significant decline-associated variants were evaluated for prior associations with relevant pulmonary traits (COPD, death from respiratory disease, cross-sectional lung function, asthma/allergic disease, emphysema/chronic bronchitis, and other/non-specified respiratory disease). Dots connected by lines depict intersecting sets and vertical bars show the number of variants in each intersection. The first column shows the variants identified in this GWAS that were not previously reported for the pulmonary traits evaluated.

Additionally, several variants reported in previous GWAS for genome-wide significant associations with cross-sectional spirometry measures (164 variants), COPD (63 variants), asthma (113 variants), and emphysema (2 variants) showed nominal evidence (p<0.05) for associations with lung function decline phenotypes in the cross-ancestry or ancestry-specific analyses (Table S7). We were unable to fully assess LD of our decline-associated variants with these previously identified variants as the majority of our variants were not present in the 1000 Genomes reference panel.

### Lung function decline is heritable and genetically correlated with relevant pulmonary traits

We estimated heritability of lung function decline in the FHS cohort accounting for sex, age at study baseline, and ever/never smoking status at 0.04 (p=0.180) for FEV_1_ decline, 0.06 (p=0.124) for FVC decline, and 0.15 (p=0.001) for FEV_1_/FVC decline. We performed analyses estimating genetic correlation of lung function decline with relevant pulmonary traits only for decline in FEV_1_/FVC (Table S8) because it was the only decline phenotype with a statistically significant heritability estimate. These analyses suggested a positive correlation of FEV_1_/FVC decline with baseline FVC (corr=0.22, se=0.11, p=0.051) and carbon monoxide diffusion capacity (corr=0.42, se=0.20, p=0.052) and a negative correlation with COPD at the last study visit (corr=-0.58, se=0.24, p=0.070) and interstitial lung abnormality (corr=-0.50, se=0.30, p=0.065). There was no evidence of genetic correlation of FEV_1_/FVC decline with baseline FEV_1_, COPD at first study visit, asthma, emphysema, or immunoglobulin E levels.

### Lung function decline associated-variants implicate 38 genes

Gene-based testing of cross-ancestry and ancestry-specific GWAS results implicated a total of 38 genes significant for at least one decline phenotype (Table S9). Comparing the Z-scores and p-values for these genes across ancestries and decline phenotypes highlighted consistency across ancestry or phenotype for several genes, including *XIRP2*, *GRIN2D*, *SATB1*, *MARCHF4*, *SIPA1L2*, *ANO5*, *H2BC10*, and *FAF2* (Figure 4A). GTEx evaluation confirmed expression of these genes in lung tissue (Figure 4B). Variants mapped to 28 of the decline-associated genes showed evidence for prior associations with spirometry or pulmonary disease traits in either the GWAS catalogue or Phenoscanner (Table S10). The remaining 10 genes, including *FAF2*, *MARCHF4*, *GRIN2D*, *AURKA*, *MAP4K2*, *H2BC10*, *COG7*, *MEN1*, *SLA2*, and *CD163*, were not previously reported to be associated with lung disease–related traits.

**Figure 4.**
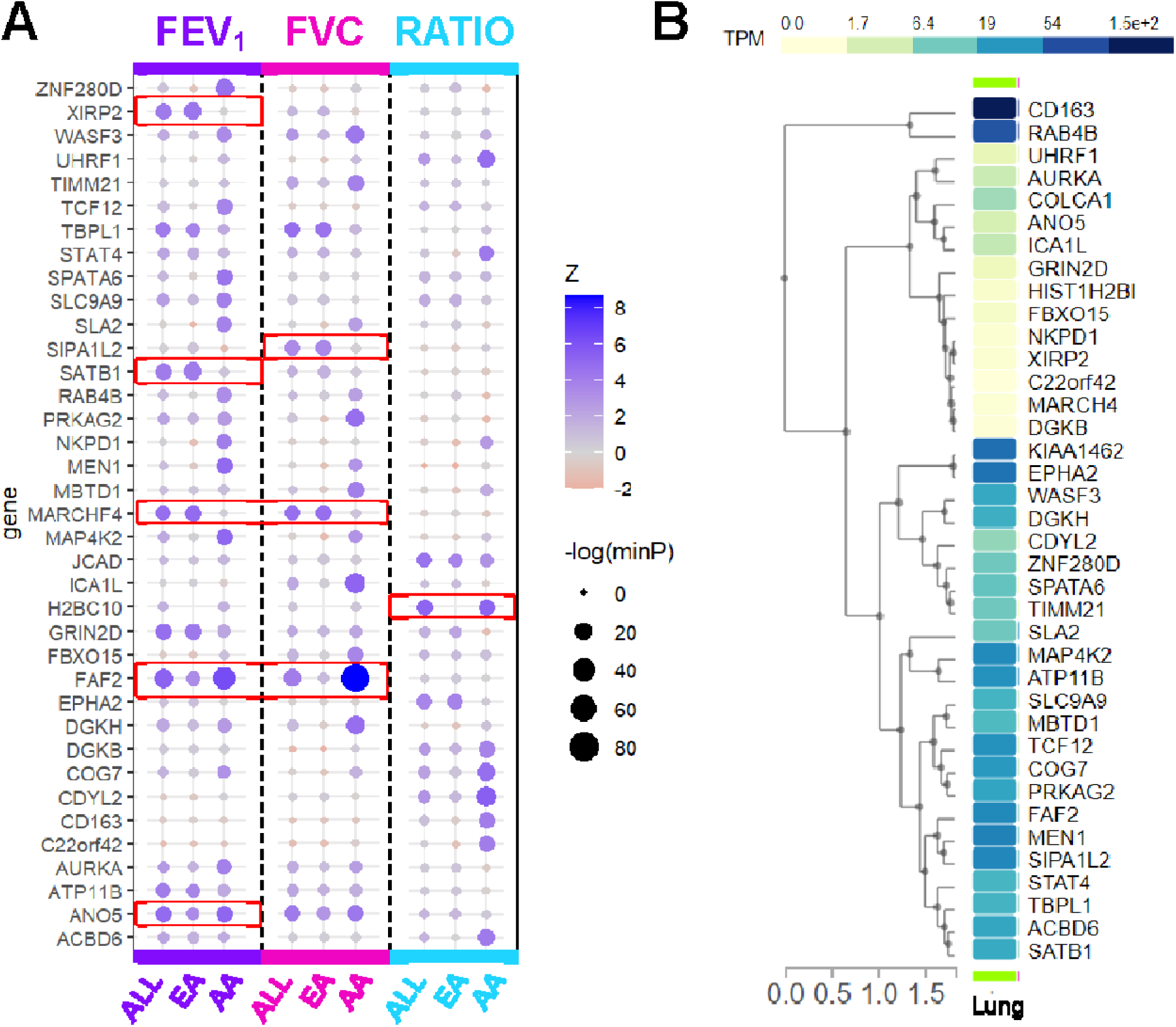
Gene-Based Testing of Cross-Ancestry and Ancestry-Specific GWAS Results. Genes implicated from gene analysis of our GWAS results. A) Z-score comparison across ancestries and decline phenotypes. Color corresponds to Z score value and circle size corresponds to p-value. Red boxes highlight genes with consistent and significant associations across ancestries or phenotypes. B) GTEx data for lung tissue. *Note: KIAA1426 synonymous for JCAD; HIST1H2BI synonymous for H2BC10*

### Lung function decline-associated genes are enriched for positional and other functional gene sets

Gene set enrichment results for positional mapping of variants identified in the cross-ancestry and ancestry-specific analyses are shown in the online supplement (Table S11). Cross-ancestry results were enriched for positional gene sets at chr6p21 for decline in FEV_1_ (p = 2.9E- 14) and FEV_1_/FVC (p = 3.0E-17) and chr3q12 for decline in FVC (p = 1.5E-07) and for the GWAS catalogue gene-set for height (p = 7.2E-08). EA results were enriched for positional gene sets at chr12q12, chr3p24, and chr7p14. AA results were enriched for 43 different positional gene sets, 16 GWAS catalogue gene sets (including gene sets for COPD and idiopathic pulmonary fibrosis), two cancer/computational gene sets, two chemical and genetic perturbation/curated gene sets, two micro-RNA targets gene sets, and one immunologic signature.

### Lung function decline-associated genes are enriched for biological pathways

FUMA-defined genomic loci for the variants identified in the cross-ancestry analysis showed significant enrichment (false discovery rate–adjusted p < 0.05) of Gene Ontology biological processes for the regulation of glucocorticoid and mineralocorticoid biosynthesis using both the binomial and hypergeometric testing methods (Figure 5). There was no further evidence for pathway or disease enrichment for genomic loci defined by the ancestry-specific analyses.

**Figure 5.**
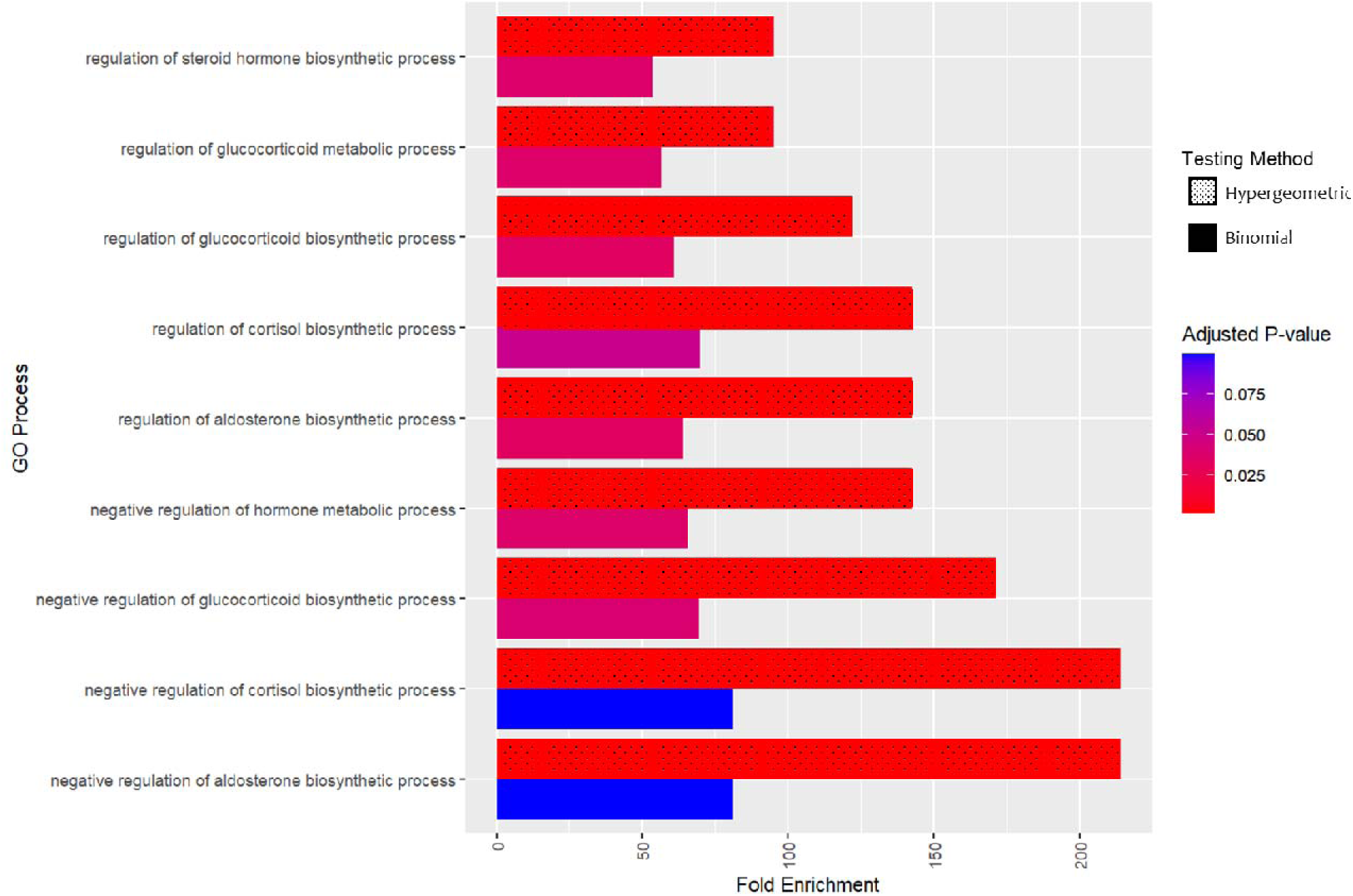
Enrichment of Processes Regulating Glucocorticoid and Mineralocorticoid Biosynthesis. Annotation class analysis of GWAS results revealed significant enrichment of Gene Ontology (GO) biological processes for corticosteroid regulatory pathways. Fold enrichment calculated using the binomial (textured) and hypergeometric (untextured) methods are shown. Color represents false discovery rate adjusted p values.

### Most lung function decline-associated genes are druggable targets

Of the 38 genes implicated in lung function decline, we found that 37 were protein coding genes that represent potential druggable targets. Queries across four drug repurposing databases identified 43 approved compounds that target eight genes of the druggable targets (*AURKA*, *CD163*, *DGKB*, *DGKH*, *EPHA2*, *GRIN2D*, *MAP4K2*, *PRKAG2*). The full set of identified compounds is available in Table S12. Inspection of the identified compounds highlighted the N-methyl-D-aspartate (NMDA) receptor GRIN2D and the receptor tyrosine kinase EphA2, which had 32 and 6 compounds, respectively. The remaining genes were targeted by one to three compounds each.

## DISCUSSION

In these multi-ancestry GWAS meta-analyses of lung function decline, we identified 361 novel genetic variants associated with lung function decline at genome-wide significance in general population cohorts, and the suggestive evidence of replication that we observed for some of these variant associations with decline in independent, COPD-enriched study populations and pleiotropy with other pulmonary phenotypes merit additional follow-up with larger sample sizes. We identified significant variants for both cross-ancestry and ancestry-specific analyses, with some signals observed across the FEV_1_, FVC, and FEV_1_/FVC decline phenotypes or across ancestry groups. Gene-based analyses similarly highlighted several genes with consistent associations across decline phenotypes or ancestries, including some genes that overlapped with genes implicated for cross-sectional lung function or other pulmonary traits and some genes that were (to our knowledge) novel. Pathway analyses revealed that the findings were enriched for genes contributing to several regulatory processes for corticosteroid biosynthesis and metabolism. Altogether, these findings contribute to our understanding of the genetic architecture of lung function decline and highlight a potential role of endogenous corticosteroid metabolism in the etiology of lung function decline.

The largest prior GWAS meta-analysis included over 27,000 EA participants from CHARGE and other cohorts did not identify any variants passing genome-wide significance in the overall study but identified one variant, rs507211 in *ME3*, that was associated with FEV_1_ decline in analyses limited to studies with three or more repeated measurements.^10^ Two other GWAS, both in approximately 6,500 Korean participants with three or more repeated measurements, each found one variant passing genome-wide significance:^5,6^ These included rs2445936 near *CEP164* associated with FEV_1_/FVC decline and rs2272402 in *SLC6A1* associated with decline in both FEV_1_ and FVC. Genome-wide significant findings for lung function decline in cohorts of participants with lung disease are similarly sparse, and findings for these cohorts are generally not validated in general population cohorts.^29–31^ The current study combined multi-ancestry populations from general population cohorts and leveraged newer imputation panels, increasing both sample size and genomic coverage. Our discovery of 361 genome-wide significant genetic associations greatly expanded the number of loci associated with longitudinal lung function. One variant identified in our AA analyses for FVC decline, rs32137 in *CTNND2*, was reported as nominally significant in a prior GWAS of lung function decline in asthmatics.^8^

Lung function decline is inherently interrelated with cross-sectional lung function, and chronic lung diseases are characterized by impaired lung function and accelerated lung function decline.^19,36–40^ The largest GWAS of cross-sectional lung function to date demonstrated strong associations of polygenic risk scores for impaired lung function with multiple chronic lung diseases, including COPD, asthma, emphysema, and chronic bronchitis, suggesting an overlap in underlying genetic architecture.^4^ In the present study, we also found evidence supporting genetic overlap between lung function decline and other pulmonary traits, including cross-sectional lung function, COPD, asthma, emphysema, chronic bronchitis, pulmonary fibrosis, and interstitial lung disease. Many of the variants identified in our GWAS were nearby (within 500 kb) variants previously associated with these traits, with the largest overlap in signals for pulmonary fibrosis, interstitial lung disease, and unspecified respiratory disease. Results from genetic correlation analysis of lung function decline and these traits, performed in a subset of EA participants from one cohort for which we had access to individual-level data, provided suggestive evidence of a genetic correlation between FEV_1_/FVC decline and baseline FVC, COPD, carbon monoxide diffusion capacity, and interstitial lung abnormality, though these findings fell just short of our nominally statistically significant threshold (p-values ranging from 0.05 to 0.07). Given the reduced sample size for this analysis, and because relatively few of our GWAS findings were from the EA-specific analyses, the heritability results may be strengthened by using individual-level data for a larger, more diverse population. Further research to understand genetic correlation of lung function decline with other pulmonary traits across different populations is warranted.

Pathway analyses of decline-associated variants highlighted several regulatory processes for corticosteroid biosynthesis and metabolism. Corticosteroids, which include glucocorticoids and mineralocorticoids, are a class of steroid hormone produced in the adrenal gland.^41^ These steroids, especially glucocorticoids, have potent anti-inflammatory and immunomodulatory effects, and synthetic versions are widely used to treat and manage various inflammatory and autoimmune diseases, including chronic lung diseases.^41,42^ Evidence also suggests that endogenous corticosteroids impact lung health throughout the lifespan. Endogenous glucocorticoids are known to play a role in lung maturation, which in turn can impact longitudinal lung function trajectories.^42^ Previous studies have demonstrated associations of low endogenous cortisol, the predominant glucocorticoid, with accelerated lung function decline.^43,44^ Endogenous cortisol levels have also been associated with risk and severity of asthma, COPD, and chronic nonspecific lung disease.^45–49^ Additionally, genetic variants in the *SERPINA6/SERPINA1* locus, which encodes proteins that impact cortisol bioavailability, have been associated with impaired lung function and are among the most established genetic risk factors for COPD.^50–53^ Our findings showing enrichment of corticosteroid regulatory pathways, including biosynthesis and metabolism of cortisol, contribute to the evidence supporting a role of corticosteroids in lung health.

Many of the genes identified in our GWAS findings have not been previously implicated in lung function. However, our drug repurposing analysis based on these genes identified compounds associated with known lung health modulators, such as the antioxidant N-acetylcysteine (NAC) that has been evaluated in COPD pathogenesis and progression^54^ and the expectorant guaifenesin that is used to thin mucus and relieve congestion in chronic lung disease,^55^ increasing our confidence that these findings could translate to actionable targets. This convergence of established and novel findings strengthens the foundation for future research endeavors.^54^

The identification of the *GRIN2D* gene, which encodes an NMDA receptor, is of particular interest, as emerging evidence is demonstrating the importance of NMDA receptors in lung function. Previous studies found that functional NMDA receptors are present in human lung tissue and lead to calcium release and airway contraction.^56^ Furthermore, NMDA receptor modulation has been proposed as part of a multi-pronged approach to improve airway smooth muscle function in Severe Acute Respiratory Syndrome patients.^57,58^ *GRIN2D*, as one of the eight genes with consistent associations across ancestries or decline phenotypes, is targeted by 32 approved compounds and represents a promising drug repurposing target. Further investigation into these compounds and the role of *GRIN2D* in lung function is needed to elucidate the specific mechanism and therapeutic potential.

This study has several limitations. First, to increase sample size and power for discovery, we combined all contributing general population cohorts rather than reserving cohorts for replication. To mitigate this and to increase clinical relevance, we replicated the analyses in two cohorts enriched for COPD. However many of the variants identified in our discovery analyses were characterized by low minor allele frequencies (MAF < 0.05), and while most variants had high imputation quality (imputation value > 0.9), increasing our confidence in these findings, our ability to replicate these variant associations was limited by the smaller sample sizes of the replication cohorts, meaning that a majority of the decline-associated variants were not available (did not pass our QC criteria) for replication analyses, especially for the AA and cross-ancestry analyses. These analyses were also limited to EA and AA participants; further research replicating our findings in larger, more diverse populations is warranted. Second, a majority of the variants identified in the AA-specific analyses only passed our QC criteria in one cohort.

Although this cohort represented nearly half the total AA participants, further investigation of these variants in cohorts with larger AA sample sizes is needed to verify these findings. Third, our multi-ancestry analyses included participants identifying as White, Black, Asian, or Hispanic, analyzed separately and combined through meta-analysis. Although multiple cohorts included both White and Black participants, only one cohort included Asian and Hispanic participants and the small sample sizes limited our ability to perform ancestry-specific analyses for these participant groups. Fourth, the length of follow-up and number of repeated measures varied across contributing cohorts, which could have contributed to heterogeneity in the precision of estimates of variant–lung function decline associations. Lastly, although we accounted for key covariates for lung function and its decline, most notably by stratifying by sex and adjusting for smoking variables, we did not evaluate interactions of these covariates with our findings. Sex-specific and gene-by-smoking interaction analyses could shed further light on genetics underlying differences in lung function decline observed by these factors.

In summary, in the first multi-ancestry and largest GWAS of lung function decline to date, we leveraged newer genetic imputation methods to achieve greater coverage of the genome and identified numerous genome-wide significant variants for decline in FEV_1_, FVC, and FEV_1_/FVC, most of which represent novel signals for lung function decline. Our results, which overlapped with previously reported genetic signals for several related pulmonary traits, implicated 38 genes and were enriched for processes involving corticosteroid biosynthesis and metabolism, underscoring existing evidence for a role of corticosteroids in chronic lung health. Our findings contribute to our understanding of the genetics underlying age-related lung function decline and highlight relevant genes, biological pathways, and potential drug targets that could be repurposed to slow decline and treat lung disease.

## Data Availability

Summary data produced in the present study are available upon reasonable request to the authors. Preexisting data access policies for each of the contributing cohort studies specify that research data requests can be submitted to each steering committee; these will be promptly reviewed for confidentiality or intellectual property restrictions and will not unreasonably be refused. Individual-level patient data may be further restricted by consent, confidentiality, or privacy laws/considerations.

## ACKNOWLEDGEMENTS

The authors thank the staff and participants of the Atherosclerosis Risk In Communities (ARIC) Study, the Coronary Artery Risk Development in Young Adults (CARDIA) Study, the Cardiovascular Health Study (CHS), the Framingham Heart Study (FHS), the Health, Aging and Body Composition (HABC) Study, the Multi-Ethnic Study of Atherosclerosis (MESA), the UK Biobank, the Genetic Epidemiology of COPD (COPDGene) Study and the Evaluation of COPD Longitudinally to Identify Predictive Surrogate End (ECLIPSE) Study for their important contributions.

## AUTHOR CONTRIBUTIONS

The study was designed and conceived by BKP, JZ, JD, PAC and DBH, with input from SHC, GO’C, SAG, AM, RGB, KN and MHC. AM, RBG, SAG, KN, ES, BY, MT and MHC provided the data and supervised the data analysis in each cohort. BKP, JZ, TMB, JC, CD, HS, NQN, JS and CT performed cohort-specific analyses and/or performed meta-analysis. BP, NCG, MM, JS, and SHC performed follow-up and downstream functionality analyses. BKP, JZ, NCG, JD, PAC and DBH interpreted the results. BKP, PAC and DBH drafted and edited the manuscript with input from all authors. All authors reviewed and contributed to the discussion of findings and the writing and/or editing of the manuscript and approved the final version.

## FOOTNOTES

### Atherosclerosis Risk in Communities

The ARIC study has been funded in whole or in part with Federal funds from the National Heart, Lung, and Blood Institute, National Institutes of Health, Department of Health and Human Services, under Contract nos. (HHSN268201700001I, HHSN268201700002I, HHSN268201700003I, HHSN268201700004I, HHSN268201700005I). Funding was also supported by R01HL087641, R01HL059367 and R01HL086694; National Human Genome Research Institute contract U01HG004402; and National Institutes of Health contract HHSN268200625226C. The authors thank the staff and participants of the ARIC study for their important contributions.

### Cardiovascular Health Study

This CHS research was supported by NHLBI contracts HHSN268201200036C, HHSN268200800007C, HHSN268201800001C, N01HC55222, N01HC85079, N01HC85080, N01HC85081, N01HC85082, N01HC85083, N01HC85086, 75N92021D00006; and NHLBI grants U01HL080295, R01HL087652, R01HL105756, R01HL103612, R01HL120393, and U01HL130114 with additional contribution from the National Institute of Neurological Disorders and Stroke (NINDS). Additional support was provided through R01AG023629 from the National Institute on Aging (NIA). A full list of principal CHS investigators and institutions can be found at CHS-NHLBI.org. The provision of genotyping data was supported in part by the National Center for Advancing Translational Sciences, CTSI grant UL1TR001881, and the National Institute of Diabetes and Digestive and Kidney Disease Diabetes Research Center (DRC) grant DK063491 to the Southern California Diabetes Endocrinology Research Center.

### Coronary Artery Risk Development in Young Adults Study

The Coronary Artery Risk Development in Young Adults Study (CARDIA) is conducted and supported by the National Heart, Lung, and Blood Institute (NHLBI) in collaboration with the University of Alabama at Birmingham (N01-HC95095 & N01-HC48047), University of Minnesota (N01-HC48048), Northwestern University (N01-HC48049), and Kaiser Foundation Research Institute (N01- HC48050).

### Framingham Heart Study

The Framingham Heart Study is conducted and supported by the National Heart, Lung, and Blood Institute (NHLBI) in collaboration with Boston University (Contract No. N01-HC-25195, HHSN268201500001I and 75N920 19D00031). Funding for SHARe Affymetrix genotyping was provided by NHLBI Contract N02-HL64278. SHARe Illumina genotyping was provided under an agreement between Illumina and Boston University. *Health, Aging and Body Composition Study:* The HABC study was supported by National Institute on Aging (NIA) Contracts N01-AG-6-2101; N01-AG-6-2103; N01-AG-6-2106; NIA grant R01-AG028050, and NINR grant R01-NR012459. This research was funded in part by the Intramural Research Program of the NIH, National Institute on Aging.

### Multi-Ethnic Study of Atherosclerosis

MESA and the MESA SHARe projects are conducted and supported by the National Heart, Lung, and Blood Institute (NHLBI) in collaboration with MESA investigators. Support for MESA is provided by contracts 75N92020D00001, HHSN268201500003I, N01-HC-95159, 75N92020D00005, N01-HC-95160, 75N92020D00002, N01-HC-95161, 75N92020D00003, N01-HC-95162, 75N92020D00006, N01-HC-95163, 75N92020D00004, N01-HC-95164, 75N92020D00007, N01-HC-95165, N01-HC-95166, N01- HC-95167, N01-HC-95168, N01-HC-95169, UL1-TR-000040, UL1-TR-001079, and UL1-TR-001420, UL1TR001881, DK063491, and R01HL105756. Funding for SHARe genotyping was provided by NHLBI Contract N02-HL-64278. Genotyping was performed at Affymetrix (Santa Clara, California, USA) and the Broad Institute of Harvard and MIT (Boston, Massachusetts, USA) using the Affymetrix Genome-Wide Human SNP Array 6.0. The authors thank the other investigators, the staff, and the participants of the MESA study for their valuable contributions. A full list of participating MESA investigators and institutes can be found at http://www.mesa-nhlbi.org.

### UK Biobank

This research has been conducted using the UK Biobank Resource under Application Number 648 using the ALICE High Performance Computing Facility at the University of Leicester. The study was supported by Wellcome Trust Awards WT202849/Z/16/Z and WT225221/Z/22/Z, and by UKRI Grant MC_PC_19004 (BREATHE Health Data Research Hub). The study was partially supported by the NIHR Leicester Biomedical Research Centre and an NIHR Senior Investigator Award to M.D.T; views expressed are those of the author(s) and not necessarily those of the NHS, the NIHR or the Department of Health. The funders had no role in the design of the study.

### Genetic Epidemiology of COPD (COPDGene)

The COPDGene study was described and supported by Award Number U01 HL089897 and Award Number U01 HL089856 from the NHLBI. The content is solely the responsibility of the authors and does not necessarily represent the official views of the National Heart, Lung, and Blood Institute or the National Institutes of Health. The COPDGene project is also supported by the COPD Foundation through contributions made to an Industry Advisory Board comprised of AstraZeneca, Boehringer Ingelheim, GlaxoSmithKline, Novartis, Pfizer, Siemens and Sunovion. The funders had no role in the design of the study. Whole genome sequencing (WGS) for COPDGene was conducted through the NHLBI-funded Trans-Omics in Precision medicine (TOPMed) program (phs000951) and was performed at the University of Washington Northwest Genomics Center (3R01 HL089856-08S1) and the Broad Institute of MIT and Harvard (HHSN268201500014C). A full list of COPDGene investigators and core units can be found at https://copdgene.org.

### Evaluation of COPD Longitudinally to Identify Predictive Surrogate End-points (ECLIPSE)

The ECLIPSE study (NCT00292552; GSK code SCO104960) was funded by GlaxoSmithKline. WGS for ECLIPSE was conducted through the NHLBI-funded Trans-Omics in Precision medicine (TOPMed) program (phs001472) and was performed at the McDonnell Genome Institute at Washington University (HHSN268201600037I). The full list of ECLIPSE investigators has been published elsewhere^1,2^.

