## Supplemental Materials for "Multi-ancestry genome-wide association study reveals novel genetic signals for lung function decline"

### Study populations and spirometry measurements

*Atherosclerosis Risk in Communities (ARIC)*: The ARIC study is a population-based longitudinal cohort of about 15,800 primarily Black and White middle-aged men and women, recruited from four communities in the U.S. between 1987 and 1989. Spirometry was performed at clinic visits 1 (1987–1989), 2 (1990–1992), and 5 (2011–2013). Collins Survey II water-sealed spirometers (Warren E. Collins Inc., Braintree, MA) were used for visits 1 and 2, and SensorMedics model 1022 dry rolling seal spirometers (OMI, Houston, TX) were used for visit 5.^1^

*Coronary Artery Risk Development in Young Adults (CARDIA):* The CARDIA study is a population-based longitudinal cohort of about 5,115 Black and White young adults aged 18 to 30, recruited from four U.S. metropolitan areas: Birmingham, AL, Chicago, IL, Minneapolis, MN, and Oakland, CA. CARDIA began in 1985–1986.^1^ Spirometry was performed at baseline (1985–1986) and exam years 2 (1987–1988), 5 (1990–1991), 10 (1995–1996) and 20 (2005–2006). Collins Survey II water-sealed spirometers were used at baseline and exam years 2, 5 and 10, and SensorMedics model 1022 dry rolling seal spirometers were used at exam year 20.^1^

*Cardiovascular Health Study (CHS)***:** CHS is a population-based longitudinal study of 520 men and women ≥ 65 years of age, recruited from four U.S. communities: Forsyth County, NC, Sacramento County, CA, Washington County, MD, and Allegheny County (Pittsburgh), PA, in 1989. An additional 687 Black men and women were recruited starting in 1992. Spirometry was performed at cohort examination 2 (1989–1990), 6 (1993–1994), 9 (1996–1997) and 18 (2005–2006). Collins Survey II water-sealed spirometers were used at exams 2, 6, and 9, and EasyOne flow-sensing spirometers (ndd Medical Technologies, Inc., Andover, MA) were used for exam 18.^1^ For this study, the exam 9 data for FVC were excluded due to concerns regarding comparability between the FVC measures at exams 6 and 9, as recommended by CHS investigators.^2^

*Framingham Heart Study (FHS)*: FHS is a household based, generational cohort in Framingham, MA, established in 1948. The Offspring cohort began in 1971 and is comprised of around 5,100 children of the original cohort and spouses of these children. The mean age at the time of enrollment was 36 years. Overall, 99.7% of all FHS participants (three generations) were self-reported White. Spirometry was performed at Offspring exams 3 (1983–1987), 5 (1991–1995), 6 (1995–1998), 7 (1998–2001), 8 (2005–2008), and 9 (2011–2014). Collins Survey II water-sealed spirometers were used for all exams.

*Health, Aging and Body Composition (HABC):* HABC is a population-based longitudinal study consisting of 3,075 Black and White men and women recruited from two US communities: Pittsburgh, PA and Memphis, TN, between March 1997 and July 1998. Participants were 70–79 years of age at recruitment. Spirometry was performed at baseline (1997–1998) and cohort examinations 5 (2001–2002, 8 (2004–2005), and 10 (2006–2007). SensorMedics model 1022 dry rolling seal spirometers were used for the baseline exam and exam 5, and EasyOne flow-sensing spirometers were used for exams 8 and 10.^1^

*Multi-Ethnic Study of Atherosclerosis (MESA):* MESA is a population-based cohort consisting of 6,814 Black, White, Hispanic, and Chinese-American men and women 45 to 84 years of age, recruited from six sites in the U.S., including St. Paul, MN, Los Angeles, CA, northern Manhattan, NY, Forsyth County, NC, Chicago, IL, and Baltimore City and County, MD, from July 2000 to August 2002. Spirometry was performed at examination 3 or 4 (2004–2007), 5 (2010–2011), and 6 (2016–2017). SensorMedics model 1022 dry rolling seal spirometers were used for all exams.^1^

*UK Biobank (UKBB):* The UKBB is a large-scale prospective study consisting of a half million UK residents, aged 40-69 at recruitment. Spirometry was performed in all participants at the initial assessment (2006–2010), and in a proportion of participants who were invited back for repeat assessments at three later time points (2012–2013, 2014+, and 2019+). Vitalograph Pneumotrac 6800 spirometers (Vitalograph Ltd., Buckingham, England) were used for all spirometry measurements.

*Evaluation of COPD Longitudinally to Identify Predictive Surrogate End-points (ECLIPSE)***:** The ECLIPSE study is a longitudinal observational study consisting of 2,164 smokers with COPD at the study baseline and a smaller number of smoking (337) and nonsmoking (245) controls without COPD at the study baseline. Recruitment occurred from December 2005-December 2006. Participants were aged 45-70 at recruitment and all participants were self-reported non-Hispanic White. Inclusion criteria for COPD cases included Global Initiative for Chronic Obstructive Lung Disease (GOLD) grades 2-4 by spirometry with post-bronchodilator FEV1 < 80% predicted and FEV1/FVC ≤ 0.7 and at least 10 pack-years of cigarette smoking. Inclusion criteria for controls included normal spirometry with post-bronchodilator FEV1>85% predicted and FEV1/FVC > 0.7. Spirometry was performed at study baseline, three months, six months and every six months thereafter for three years.^3,4^

*Genetic Epidemiology of Chronic Obstructive Pulmonary Disease (COPDGene)***:** The COPDGene study is an ongoing prospective observational study consisting of more than 10,000 smokers with and without COPD and a smaller number of non-smoking controls recruited from 21 clinical sites across the United States. Inclusion criteria included at least 10 pack-years of cigarette smoking (except for non-smoking controls) and self-identification as non-Hispanic white or African American, with planned enrollment of 2/3 non-Hispanic White and 1/3 African American. Spirometry was performed at the study baseline and the 5 and 10 year follow-up visits using the ndd EasyOne Spirometer (Zurich, Switzerland).^5,6^

### Statistical analysis

Associations of genetic variants with lung function decline were evaluated in each cohort using general estimating equations with robust standard error. Repeated measurements of FEV_1_, FVC, or FEV_1_/FVC were regressed on variant, elapsed time since first lung function measurement, and the variant × elapsed time multiplicative interaction term. Analyses were stratified by sex and self-reported race/ethnicity (as a proxy for genetic ancestry), and adjusted for covariates selected *a priori* based on prior knowledge of lung function predictors. These included baseline age, height, weight (FVC only), baseline ever smoke history, current smoking status, baseline smoking pack-years, cigarettes per day, spirometry protocol (dummy variable indicating a change in spirometry protocol between adjacent PFT measurements), study site (for multi-site cohorts) and genotype principal components. Median-centered squared terms for age and height were also included to account for non-linear effects of age and height on lung function parameters. Weight was included in the FVC model only based on its stronger association with a restrictive (FVC) vs obstructive (FEV_1_, FEV_1_/FVC) phenotype.

**REFERENCES**

1.	Oelsner, E.C., Balte, P.P., Cassano, P.A., Couper, D., Enright, P.L., Folsom, A.R., Hankinson, J., Jacobs, D.R., Kalhan, R., Kaplan, R., et al. Harmonization of Respiratory Data From 9 US Population-Based CohortsThe NHLBI Pooled Cohorts Study. Am. J. Epidemiol. https://doi.org/10.1093/aje/kwy139.

2.	Spirometry Comparability | chs-nhlbi https://chs-nhlbi.org/internal/SpirometryComparability.

3.	Hurst, J.R., Vestbo, J., Anzueto, A., Locantore, N., Müllerova, H., Tal-Singer, R., Miller, B., Lomas, D.A., Agusti, A., MacNee, W., et al. (2010). Susceptibility to Exacerbation in Chronic Obstructive Pulmonary Disease. N. Engl. J. Med. *363*, 1128–1138. https://doi.org/10.1056/NEJMoa0909883.

4.	Vestbo, J., Anderson, W., Coxson, H.O., Crim, C., Dawber, F., Edwards, L., Hagan, G., Knobil, K., Lomas, D.A., MacNee, W., et al. (2008). Evaluation of COPD Longitudinally to Identify Predictive Surrogate End-points (ECLIPSE). Eur. Respir. J. *31*, 869–873. https://doi.org/10.1183/09031936.00111707.

5.	Regan, E.A., Hokanson, J.E., Murphy, J.R., Make, B., Lynch, D.A., Beaty, T.H., Curran-Everett, D., Silverman, E.K., and Crapo, J.D. (2010). Genetic Epidemiology of COPD (COPDGene) Study Design. COPD *7*, 32–43. https://doi.org/10.3109/15412550903499522.

6.	Maselli, D.J., Bhatt, S.P., Anzueto, A., Bowler, R.P., DeMeo, D.L., Diaz, A.A., Dransfield, M.T., Fawzy, A., Foreman, M.G., Hanania, N.A., et al. (2019). Clinical Epidemiology of COPD: Insights From 10 Years of the COPDGene Study. Chest *156*, 228–238. https://doi.org/10.1016/j.chest.2019.04.135.

**Table S1) Genotype and imputation details for each study**

| Cohort | Ancestry groups | Genotyping platform | QC steps and specific thresholds used for excluding genotyped variants |
| --- | --- | --- | --- |
| ARIC | EA, AA | Affymetrix 6.0 | call rate<95%, HWE P<10^6^, MAF<1%, or no chromosomal location |
| CARDIA | EA, AA | Affymetrix 6.0 | call rate < 95%, HWE P < 10-5, duplicates, monomorphic variants, MAF < 1% (AA only) |
| CHS | EA, AA | Illumina 370CNV BeadChip (EA), Illumina HumanOmni1-Quad_v1 BeadChip (AA) | Call rate <97%, HWE P < 10-5, >2 duplicate errors or Mendelian inconsistencies. *At the time of analysis, variants were excluded for variance on the allele dosage <=0.01. |
| FHS | EA | Affymetrix 500K + 50K Human Gene Focused Panel | call rate < 96.9%, HWE P < 10-6, MAF < 1%, Mendelian errors > 1000, not being on chromosomes 1–22 or X, duplicates |
| HABC | EA, AA | Illumina Human1M-Duo | call rate < 95%, HWE P < 10^-6^, or MAF < 1% |
| MESA | EA, AA, HA, CHN | Affymetrix 6.0 | call rate < 95%, heterozygosity >53%, or monomorphic variants |
| UKBB | EA | Affymetrix UK Biobank or UK BiLEVE Axiom Arrays |  |

Figure S1) Quantile-quantile plots and genomic inflation values for cross-ancestry and ancestry-specific meta-analyses. The plots compare the observed vs. expected *P* values for testing of the variant by time interaction term in relation to FEV_1_, FVC or FEV_1_/FVC. The corresponding genomic inflation factors are shown, as calculated across all variants before the exclusion of previously implicated variants.

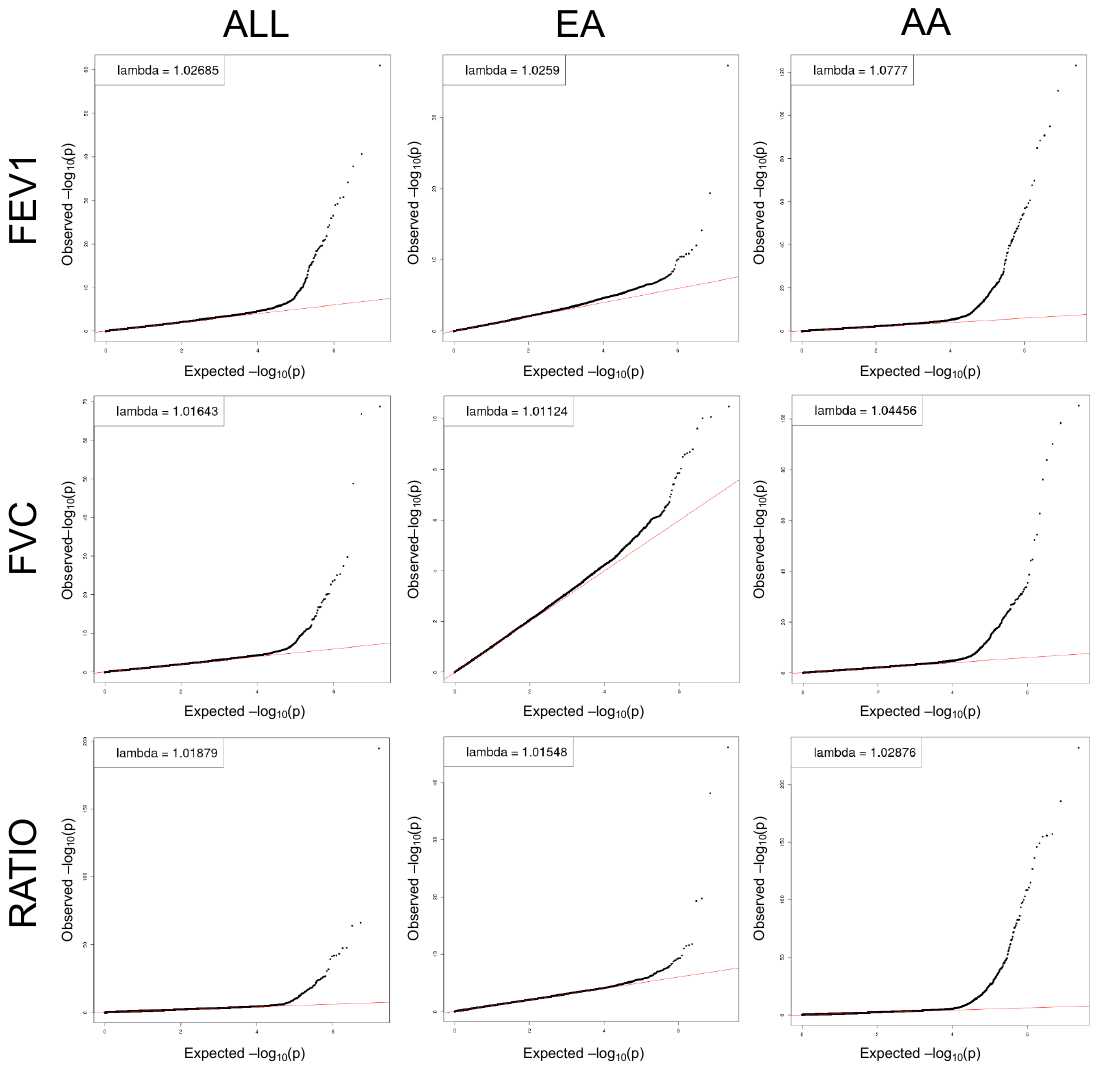

Figure S2) Violin plot of minor allele frequency (MAF) distributions of variants with p<5E-08 across decline phenotypes and ancestry. The plot shows density curves for the log_10_ transformed MAF distribution of significant variants identified from the cross-ancestry and ancestry specific analyses for decline in FEV_1_, FVC and FEV_1_/FVC. The dotted red line represents MAF of 0.05.

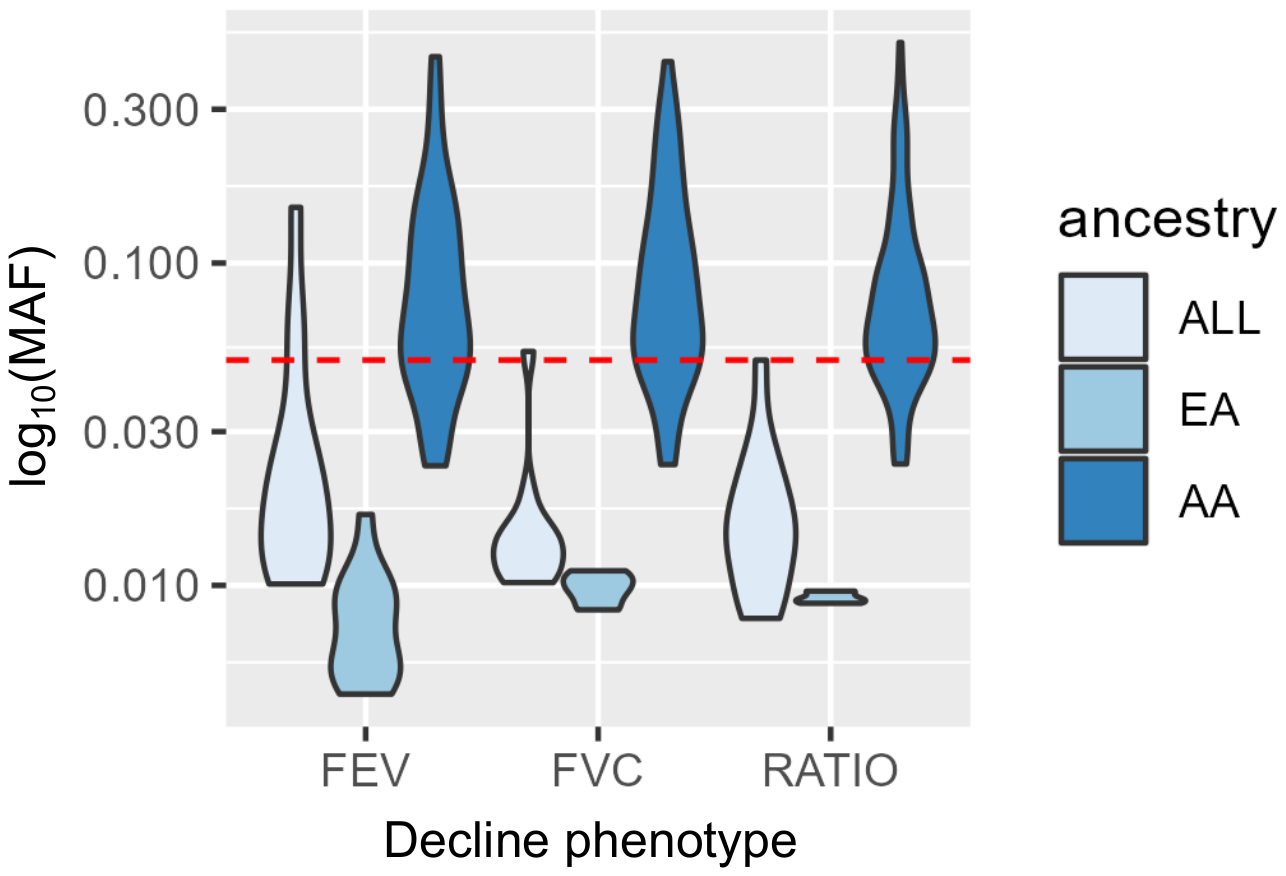

Figure S3) Circular Manhattan plot for European Ancestry analysis. Genome-wide results for decline in FEV_1_ (outer circle), FVC (middle circle), and FEV_1_/FVC (inner circle) from the European ancestry analyses. Dotted red lines denote the genome-wide significance threshold of p = 5E-08.  Red circles represent variants passing genome-wide significance.

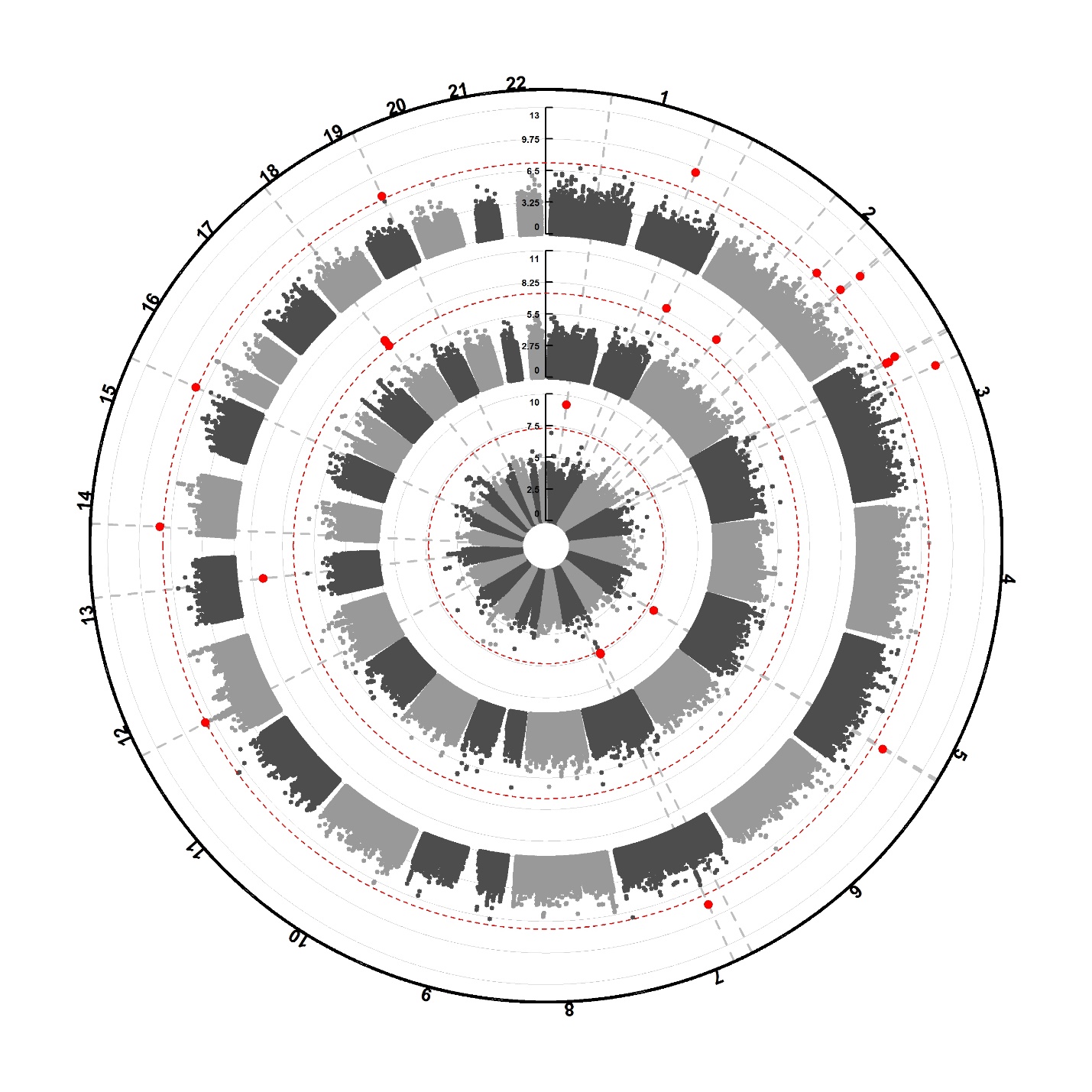

**FEV_1_/FVC**

**FVC**

**FEV1**

Figure S4) Circular Manhattan plot for African ancestry analyses. Genome-wide results for decline in FEV_1_ (outer circle), FVC (middle circle), and FEV_1_/FVC (inner circle) from the African ancestry analyses. Dotted red lines denote the genome-wide significance threshold of p = 5E-08.  Red circles represent variants passing genome-wide significance.

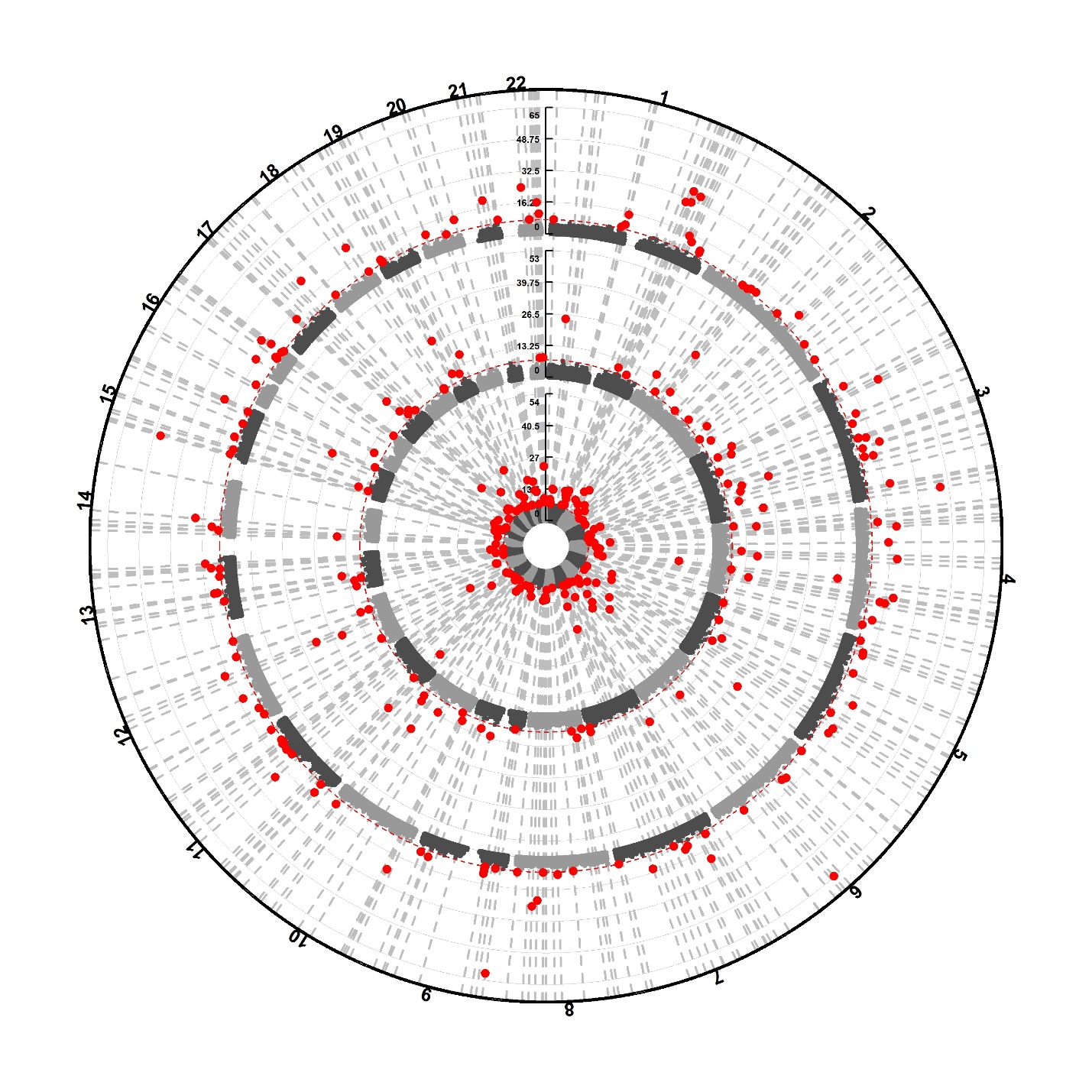

**FEV_1_**

**FEV_1_/FVC**

**FVC**
